# Autoregressive With Exogenous Input (ARX) Decision Support for Blood Pressure Maintenance During Cesarean Delivery Under Spinal Anesthesia: A Prospective Pilot Study With Matched Nonconcurrent Controls

**DOI:** 10.64898/2026.02.03.26345255

**Authors:** Vesela P. Kovacheva, Niharika Mahesh, Sherwin C. Davoud, Ricardo Kleinlein, Nolan Wheeler, Prakhar Kapoor, Bernard Rosner, Basak Ozaslan, Eleonora M. Aiello, Francis J. Doyle

## Abstract

**Background:** Spinal anesthesia for cesarean delivery commonly causes maternal hypotension, which may compromise uteroplacental perfusion and maternal comfort. Guidelines recommend maintaining maternal blood pressure near baseline with prophylactic vasopressor strategies, yet titration remains reactive. We evaluated an autoregressive with exogenous input (ARX) decision-support algorithm that provides real-time forecasts of maternal mean arterial pressure (MAP) to support vasopressor management during cesarean delivery under spinal anesthesia.

**Methods:** In this single-center, open-label, prospective pilot study, 20 pregnant patients at term undergoing elective cesarean delivery under spinal anesthesia received standard care supplemented by ARX-generated MAP predictions at 1-, 2- and 3-minute horizons. Clinicians titrated phenylephrine per institutional protocol while reviewing ARX predictions, retaining full autonomy for dosing decisions. Predictive performance was quantified using root mean square error (RMSE), mean absolute error (MAE), coefficient of determination (R²), and fraction of improvement in total error (FIT). ARX-guided patients were matched 1:2 to nonconcurrent controls (n = 40) on attending anesthesiologist and intrathecal bupivacaine dose, with nearest-neighbor matching on age and body mass index. Exploratory outcomes included hypotension (MAP <80% of baseline), phenylephrine dose, maternal nausea, and neonatal outcomes. For minute-level hypotension classification performance, sensitivity/specificity (and related metrics) were estimated using generalized estimating equations (GEE) to account for within-patient clustering of repeated observations.

**Results:** One-minute-ahead ARX predictions achieved a mean (±SD) RMSE of 3.71 ± 3.26 mmHg and MAE of 2.75 ± 2.52 mmHg, with R² 0.34 ± 0.63 and FIT 21.1% ± 18.7%. Predictive performance decreased at longer horizons. For hypotension prediction, one-minute-ahead GEE-estimated population-average sensitivity and specificity were 57.39% and 99.74%, respectively. During the observation window, in exploratory comparisons with matched nonconcurrent controls, ARX-guided patients had a shorter duration of hypotension (0.8 ± 1.9 vs 3.0 ± 3.8 minutes; P = .005) and a lower incidence of hypotension (25.0% vs 52.5%; P = .033), but a higher phenylephrine dose (1823 ± 659 vs 974 ± 328 µg; P = .001). Maternal nausea incidence was lower in the ARX group compared with matched nonconcurrent controls (5% vs 35%; P = .014), with similar neonatal outcomes.

**Conclusions:** In this prospective pilot study, an ARX decision-support algorithm provided accurate 1-minute-ahead MAP forecasts and was associated with higher phenylephrine dosing and shorter maternal hypotension duration compared with matched nonconcurrent controls. These findings support further evaluation in larger, randomized trials.

**Summary statement:** In this prospective pilot study of 20 patients undergoing cesarean delivery under spinal anesthesia, an autoregressive with exogenous input (ARX) decision-support algorithm provided real-time blood pressure forecasts and was associated with a shorter hypotension duration but higher phenylephrine dose compared with matched nonconcurrent controls. These preliminary data support further evaluation of ARX-guided, algorithmic vasopressor management in larger, multicenter trials.

**Key Points:**

- **Question:** In pregnant patients at term undergoing elective cesarean delivery under spinal anesthesia, can a real-time ARX algorithm accurately forecast MAP and support vasopressor management?
- **Findings:** One-minute-ahead forecasts were accurate (RMSE 3.71 mmHg), and ARX-guided care was associated with a shorter duration of hypotension and a higher phenylephrine dose versus matched nonconcurrent controls
- **Meaning:** Real-time MAP forecasting is feasible and warrants randomized evaluation to confirm clinical benefit and characterize trade-offs.

## INTRODUCTION

Spinal anesthesia is preferred for elective cesarean delivery, but sympathetic blockade can lead to abrupt vasodilation and maternal hypotension. Without effective prophylaxis, hypotension has been reported in up to 80% of patients.[1,2] Maternal hypotension is associated with nausea and vomiting and, when severe or prolonged, may impair uteroplacental perfusion and fetal acid–base status.[2,3]

International guidelines recommend baseline-referenced blood pressure targets and prophylactic vasopressors.[4] Phenylephrine is used as first-line therapy because it effectively maintains arterial pressure and is associated with more favorable fetal acid–base profiles than ephedrine.[2,3] Prophylactic phenylephrine infusions reduce hypotension and associated nausea and vomiting compared with reactive bolus-only approaches.[2,5] However, vasopressor dose–response varies between individuals, and higher phenylephrine doses may cause bradycardia, reactive hypertension, or reductions in maternal cardiac output that warrant careful quantification in any strategy that promotes more proactive dosing.[2,6]

Even with prophylaxis, vasopressor titration remains reactive because intermittent noninvasive blood pressure measurements may lag rapid post-spinal hemodynamic changes. Closed-loop vasopressor systems can improve blood pressure control and reduce clinician workload, but they require real-time device and infusion-pump integration.[7–9]

Forecasting-based decision support could enable proactive therapy without automated control. In non-obstetric surgery, waveform-based algorithms can predict hypotension, and early warning systems can reduce hypotension burden.[10,11] The Hypotension Prediction Index has also been evaluated in awake cesarean delivery patients under spinal anesthesia using noninvasive arterial pressure waveforms.[12] However, waveform-based approaches require continuous pressure signals and do not incorporate vasopressor dosing or intrathecal local anesthetic effects. We previously developed an interpretable autoregressive model with exogenous inputs (ARX) using mean arterial pressure (MAP), phenylephrine, and intrathecal bupivacaine dosing to generate short-horizon MAP forecasts.[1]

In this prospective pilot study, we evaluated real-time ARX-generated MAP forecasts at 1–3-minute horizons displayed to clinicians during elective cesarean delivery under spinal anesthesia. The primary objective was to quantify prospective prediction accuracy; secondary objectives included evaluation of short-horizon hypotension prediction and exploratory comparisons of hemodynamics, maternal and neonatal outcomes with matched nonconcurrent controls. We hypothesized that 1-minute-ahead ARX forecasts would provide actionable lead time and could reduce hypotension burden during the prespecified observation window (from spinal anesthesia administration until delivery or 20 minutes, whichever occurred first), potentially at the expense of increased phenylephrine exposure.

## MATERIALS AND METHODS

### Study Design

This single-center, open-label, prospective pilot study at Brigham and Women’s Hospital (Boston, MA, USA) evaluated a clinician-facing ARX decision-support display that provided real-time maternal MAP forecasts during elective cesarean delivery under spinal anesthesia. The protocol was approved by the Mass General Brigham Institutional Review Board (IRB #2025P000588). A separate IRB protocol covered the use of data for nonconcurrent matched control comparisons (IRB #2023P000178). All participants (ARX-guided patients and controls) provided written informed consent. This manuscript adheres to the applicable TREND reporting guidelines for nonrandomized evaluations of interventions.

The primary objective was to quantify prospective prediction accuracy of ARX-generated MAP forecasts at 1-, 2-, and 3-minute horizons after spinal anesthesia. Prespecified secondary objectives were to evaluate short-horizon hypotension prediction, describe system implementation, and perform exploratory comparisons of maternal and neonatal outcomes with matched nonconcurrent controls (not intended for causal inference).

### Participants

Eligible participants were adults (age ≥18 years) scheduled for elective cesarean delivery at term (≥37 weeks’ gestation) at our tertiary care hospital. Inclusion criteria were singleton pregnancy, American Society of Anesthesiologists physical status II–III, planned spinal anesthesia using hyperbaric bupivacaine, planned prophylactic phenylephrine administration, and ability to provide written informed consent.

Exclusion criteria were hypertensive disorders of pregnancy (e.g., preeclampsia), clinically significant cardiac disease or arrhythmias, planned use of vasopressors other than phenylephrine, allergy or contraindication to bupivacaine or phenylephrine, urgent or emergent cesarean delivery, and inability to cooperate with study procedures. Patients meeting any exclusion criterion were not approached for consent.

### ARX Model Description

The ARX model used in this study had been previously developed and validated using retrospective data from 172 patients who underwent cesarean delivery under spinal anesthesia at our institution.[1] In brief, a third-order ARX model was selected based on predictive accuracy, interpretability, and sensitivity across training and validation datasets. During development, performance was assessed using root mean square error (RMSE) and fraction of improvement in total error (FIT). Internal (30% hold-out, n = 52) and external (n = 84) validation demonstrated approximately 4 mmHg RMSE for 1-minute-ahead MAP forecasts.[1]

The final ARX model incorporated three key inputs: (1) past MAP measurements, (2) phenylephrine infusion and bolus dosing, and (3) intrathecal bupivacaine dose. The model operated at a one-minute sampling interval, aligned with the automated noninvasive blood pressure measurements and time-stamped vasopressor dosing records. FIT was defined as the percentage reduction in prediction error (sum of squared residuals) relative to the total sum of squared deviations of the reference signal from its mean, with higher FIT values indicating better model predictive performance. Model parameters were fixed based on the prior development work and were not re-estimated during the prospective pilot. Full details of the model structure and training procedures are published.[1] Real-time implementation details and the clinician-facing display are provided in Supplemental Digital Content (SDC) 1.

For the prospective pilot, the ARX model generated real-time forecasts of maternal MAP at prediction horizons of 1, 2, and 3 minutes. Model calculations were performed on a dedicated workstation in real time, and predictions were displayed to the anesthesia team. The ARX display did not provide dosing recommendations; it displayed predicted MAP trajectories conditional on the most recent MAP and bupivacaine and phenylephrine dosing inputs.

### Intervention and Data Collection

Spinal anesthesia was performed in the sitting or lateral position using a 25-gauge Whitacre spinal needle at the L3–L4 or L4–L5 interspace. The intrathecal solution consisted of hyperbaric bupivacaine 0.75% (12-15 mg), fentanyl 10–15 µg, and morphine 100–200 µg, according to established institutional practice. After intrathecal injection, patients were positioned supine with left uterine displacement.

Standard monitoring included noninvasive blood pressure (NIBP), electrocardiography, and pulse oximetry (GE Healthcare, Chicago, IL, USA). NIBP was recorded at one-minute intervals throughout the case. Baseline MAP was defined as the mean of the last three NIBP measurements obtained prior to spinal anesthesia administration.[13,14]

Phenylephrine was administered according to institutional protocol via continuous infusion started at 20-40 µg/min and/or intermittent bolus dosing of 80-120 µg to maintain MAP ≥80% of baseline. The institutional phenylephrine protocol is provided (Supplemental Digital Content 2). Infusion rates and bolus doses were at the discretion of the attending anesthesiologist. The ARX system did not issue explicit dosing orders; instead, it displayed real-time MAP forecasts at 1-, 2-, and 3-minute horizons on a dedicated screen adjacent to the standard anesthesia workstation. Anesthesiologists could use these predictions to inform vasopressor titration but retained full autonomy over all clinical decisions.

The ARX system was activated immediately after spinal anesthesia placement and remained active until the delivery of the neonate or 20 minutes after spinal anesthesia, whichever occurred first. A 20-minute cutoff was chosen to approximate average times from spinal anesthesia administration until delivery at our institution and standardize comparisons. Model inputs (MAP, bupivacaine, and phenylephrine dosing) were entered in real time by in-room research staff at 1-minute intervals using the study interface (Supplemental Digital Content 1). For each case, the system generated and stored minute-by-minute values for the ARX-predicted MAP at each horizon. Instances in which the attending anesthesiologist chose to act contrary to the ARX predictions were documented as “overrides,” along with the primary reason. An override was defined a priori as an explicit clinician-reported deviation from the intended vasopressor adjustment attributable to disagreement with the displayed forecast.

Intraoperative data included minute-by-minute MAP, heart rate, phenylephrine infusion rates and bolus doses, ARX predictions at each time point, and anesthesiologist overrides with reasons. For both ARX-guided cases and matched nonconcurrent controls, a trained research staff member was present in the operating room and recorded time-stamped medication dosing, maternal nausea (assessed by direct questioning, with onset and resolution times recorded), Apgar scores at 1 and 5 minutes, and NICU admission status. These outcomes were prespecified exploratory endpoints given the pilot design and limited sample size.

### Control Comparisons

To provide clinical context for the intraoperative outcomes observed in the ARX-guided cohort, we identified matched nonconcurrent controls from patients who underwent elective cesarean delivery under spinal anesthesia at our institution during the period preceding the ARX intervention (May 2024 - May 2025). These control cases were drawn from a previous prospectively collected cohort, in which trained research staff were present in the operating room and recorded time-stamped medication administration, maternal and neonatal outcomes. These patients received standard institutional management of spinal hypotension, met the same inclusion and exclusion criteria and had no exposure to ARX predictions. Baseline MAP definition, hypotension threshold, and the prespecified observation window (spinal-to-delivery or 20 minutes) were identical to those used in the ARX-guided cohort.

Matching was prespecified and performed using a deterministic greedy algorithm implemented in Python (version 3.11; Python Software Foundation). Each ARX-guided patient was required to have exact matches on two primary variables: attending anesthesiologist, to account for provider-specific differences in vasopressor management, and intrathecal bupivacaine dose category (12.0, 12.7, 13.5, 14.2, or 15.0 mg). Only nonconcurrent controls with exact matches on both variables were eligible for further consideration.

Among eligible controls, secondary matching was performed using a nearest-neighbor greedy rule based on age and body mass index (BMI). Candidate controls were ranked according to the sum of absolute differences in age and BMI, and the two best matches (smallest combined difference) were selected for each ARX-guided patient, yielding a target 1:2 matching ratio. Once a control was matched, it was removed from the pool to avoid reuse and ensure independence of matched sets. No caliper was applied.

Balance between ARX-guided patients and matched nonconcurrent controls was evaluated using standardized mean differences (SMDs) for age and BMI. For this feasibility pilot, we prespecified SMD <0.20 as an acceptable balance threshold.

### Endpoints and Measures

Hemodynamic, phenylephrine, and maternal nausea outcomes were assessed from spinal anesthesia administration until delivery or 20 minutes, whichever occurred first. Neonatal outcomes (Apgar scores at 1 and 5 minutes and NICU admission) were recorded after delivery.

The primary outcome was ARX predictive accuracy, measured as the RMSE between ARX-predicted and observed MAP at 1-, 2-, and 3-minute prediction horizons. Additional accuracy metrics included mean absolute error (MAE), coefficient of determination (R²), and FIT relative to the patient-specific total sum of squared deviations of the reference signal from its mean. Prespecified secondary outcomes included hypotension prediction performance and system implementation. Exploratory clinical outcomes compared ARX-guided patients with matched nonconcurrent controls. Hypotension was defined as MAP <80% of baseline according to our institutional practice (baseline = mean of the final three pre-spinal MAP measurements). A predicted hypotension event at time t + k was defined as ARX-predicted MAP at horizon k falling below 80% of baseline. At each prediction horizon, we evaluated sensitivity, specificity, positive predictive value (PPV), and negative predictive value (NPV) for hypotension prediction.

Exploratory maternal outcomes (ARX-guided versus matched nonconcurrent controls) measured during the spinal-to-delivery (or 20-minute window) included incidence of hypotension (defined as at least one MAP measurement <80% of baseline); duration of hypotension (cumulative minutes with MAP <80% of baseline); total phenylephrine dose (µg); number of phenylephrine boluses; number of phenylephrine infusion rate adjustments; lowest intraoperative MAP; and maternal nausea, assessed by in-room research staff via direct questioning after spinal anesthesia. Total phenylephrine dose was calculated as the sum of all bolus doses plus the infusion rate integrated over time during the prespecified observation window. Exploratory neonatal outcomes collected after delivery included Apgar scores at 1 and 5 minutes and NICU admission.

Implementation was assessed descriptively by the frequency of clinician-reported overrides, overrides per case, and recorded reasons for overrides. Because override documentation captures only explicit clinician disagreement with the forecast, it was not intended to quantify clinician engagement or reliance on the display.

All comparative analyses involving matched nonconcurrent controls were prespecified as exploratory and intended to contextualize the magnitude and variability of clinical outcomes rather than to support causal inference.

### Statistical Analysis

The sample size (n = 20) was selected to assess the feasibility of real-time deployment and to estimate predictive performance with reasonable precision. Exploratory 1:2 matching to nonconcurrent controls was planned to contextualize hemodynamic and clinical outcomes and to generate preliminary effect-size estimates for future randomized trials.

Prediction metrics (RMSE, MAE, R², and FIT) were calculated at each prediction horizon (1, 2, and 3 minutes) using all eligible one-minute intraoperative observations during the ARX-active period from spinal anesthesia placement to delivery (or 20 minutes, whichever occurred first). To avoid overweighting patients with longer spinal-to-delivery times, we computed each metric at the patient level and summarized across patients (mean ± SD).

For the hypotension prediction task, sensitivity, specificity, PPV, and NPV (and 95% CIs) were estimated at each prediction horizon using generalized estimating equations (GEE) with a binomial family and a working exchangeable correlation structure to account for within-patient clustering arising from repeated binary measurements. This population-average specification assumes a constant correlation among observations within the same patient and is appropriate when the primary interest lies in population-average performance metrics rather than in modeling fine-scale temporal dependence. Robust (sandwich) standard errors were used to obtain 95% confidence intervals. We also report the number of hypotension-positive and hypotension-negative time points to contextualize classification performance.

Clinical outcome comparisons between ARX-guided patients and matched nonconcurrent controls were conducted for contextual interpretation. Continuous outcomes (e.g., duration of hypotension, phenylephrine dose, lowest MAP) were summarized as mean ± standard deviation (SD) or median (interquartile range [IQR]), depending on distributional characteristics. Because controls were matched 1:2 to each ARX-guided patient, we summarized each matched set as the ARX-guided value and the mean of its two matched nonconcurrent controls and then compared within-set differences using paired t tests for approximately normally distributed outcomes and Wilcoxon signed-rank tests otherwise. Categorical outcomes (e.g., incidence of hypotension, maternal nausea, NICU admission) were summarized as counts and percentages and compared using the Cochran-Mantel-Haenszel test with the matched set as the stratum.

Prediction accuracy analyses included all ARX-guided patients with at least 10 one-minute epochs of complete hemodynamic and phenylephrine dosing data after spinal anesthesia or until delivery if delivery occurred earlier. Missing MAP values in the control group were rare (0.58% ± 2.46% of minute-level MAP measurements per patient) and were primarily due to transient NIBP cuff failures. By design, the interface used to supply inputs to the model for the study group did not permit missing values; instead, it automatically carried forward the last observed MAP value. For transient NIBP cuff failures in data from the control group, missing values were imputed by interpolating between the nearest preceding and subsequent measurements (linear interpolation).

All analyses were conducted using Python (version 3.11). Given the pilot design, modest sample size, and the use of matched nonconcurrent controls, statistical significance testing was considered exploratory. P values are reported for exploratory hypothesis generation, and 95% confidence intervals (CIs) are provided where feasible. All tests were two-sided with an alpha level of .05.

## RESULTS

### Study Population

Between June 16, 2025, and July 16, 2025, 20 patients undergoing elective cesarean delivery under spinal anesthesia were enrolled in the ARX-guided intervention cohort. All 20 had complete intraoperative ARX predictions and hemodynamic data suitable for inclusion in the primary predictive-accuracy analyses.

Each ARX-guided patient was matched to two nonconcurrent controls, yielding 40 matched nonconcurrent controls without ARX predictions. Exact matching on attending anesthesiologist and intrathecal bupivacaine dose category was achieved for all included pairs. Standardized mean differences for age and BMI were <0.20, consistent with the prespecified balance criterion for this feasibility pilot. Baseline demographic, obstetric, and anesthetic characteristics were broadly similar between groups (Table 1). A participant flow diagram is provided in Figure 1.

**Table 1.** Baseline characteristics of ARX-guided patients and matched nonconcurrent controls.

| Category | ARX Study<br>(n = 20) | Matched Nonconcurrent Controls<br>(n = 40) |
| --- | --- | --- |
| Age, years (mean ± SD) | 34.2 ± 5.0 | 34.9 ± 3.3 |
| Race |  |  |
| White | 14 (70.0%) | 31 (77.5%) |
| Black | 2 (10.0%) | 7 (17.5%) |
| Asian | 1 (5.0%) | 0 (0.0%) |
| Other | 3 (15.0%) | 2 (5.0%) |
| Ethnicity |  |  |
| Hispanic | 4 (20.0%) | 1 (2.5%) |
| Not Hispanic | 16 (80.0%) | 39 (97.5%) |
| Parity (median, IQR) | 1 (1-2) | 1 (1-1) |
| Gravidity (median, IQR) | 3 (2-4) | 2 (2-4) |
| Newborn gestational age, wk (mean ± SD) | 39.0 ± 0.8 | 38.6 ± 1.1 |
| Newborn weight, kg (mean ± SD) | 3.46 ± 0.6 | 3.35 ± 0.5 |
| BMI, kg/m<sup>2</sup> (mean ± SD) | 31.0 ± 8.6 | 31.8 ± 4.9 |
| Intrathecal bupivacaine, mg (median, IQR) | 13.5 (12.0-13.5) | 13.5 (12.0-13.5) |

Matching was exact on attending anesthesiologist and intrathecal bupivacaine category, with nearest-neighbor matching on age and BMI.
Abbreviations: BMI, body mass index; IQR, interquartile range; SD, standard deviation.

**Figure 1.**
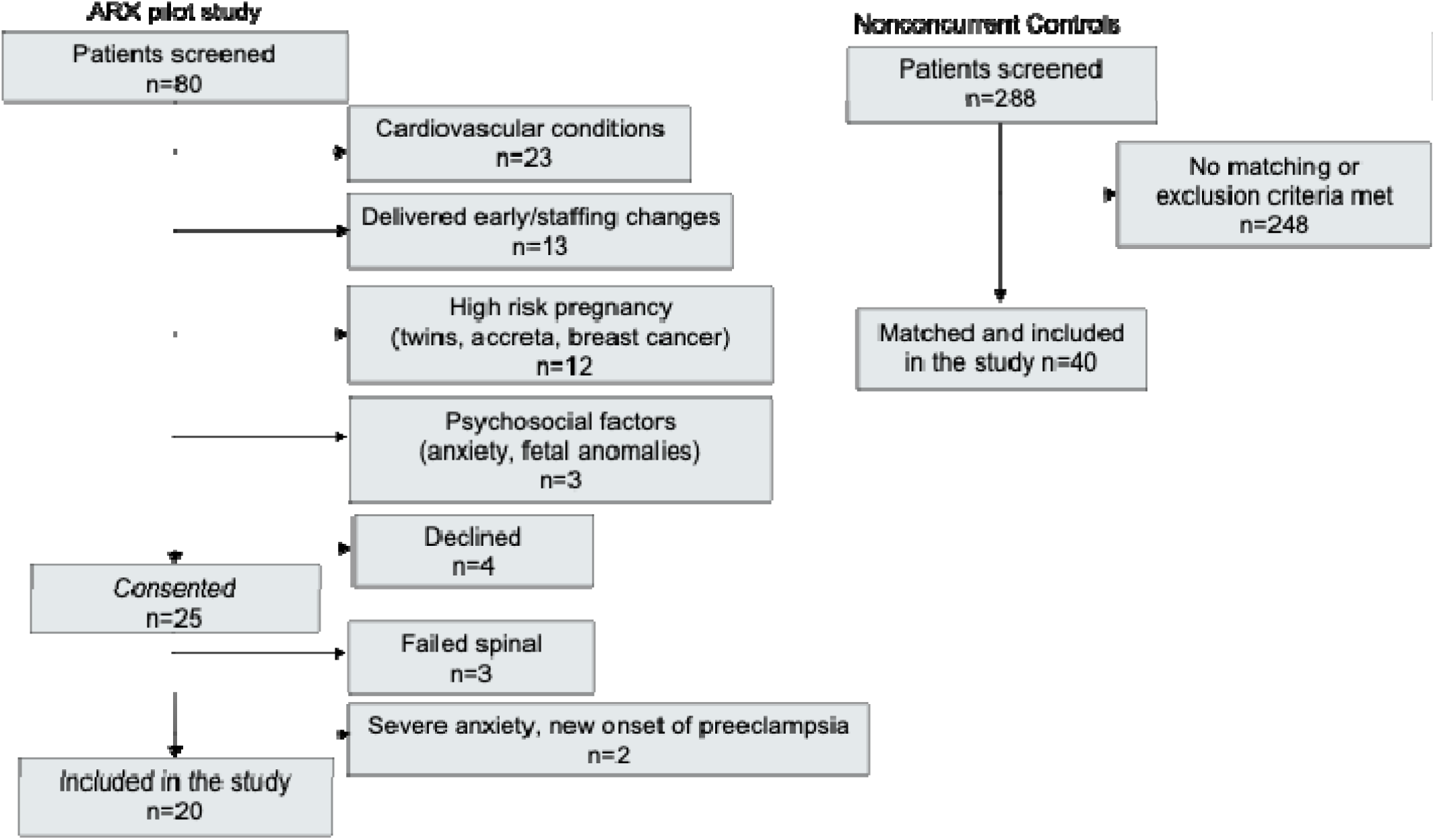
Participant flow for the ARX-guided cohort and identification/matching of nonconcurrent controls.

### ARX Predictive Accuracy

The ARX model generated real-time forecasts of maternal MAP at 1-, 2-, and 3-minute horizons for all 20 intervention cases from spinal anesthesia administration until delivery or 20 minutes, whichever occurred first. Across the cohort, 398 one-minute observations were available for 1-minute-ahead evaluation, 378 for 2-minute-ahead evaluation, and 358 for 3-minute-ahead evaluation. Predictive performance metrics are summarized in Table 2.

**Table 2.**
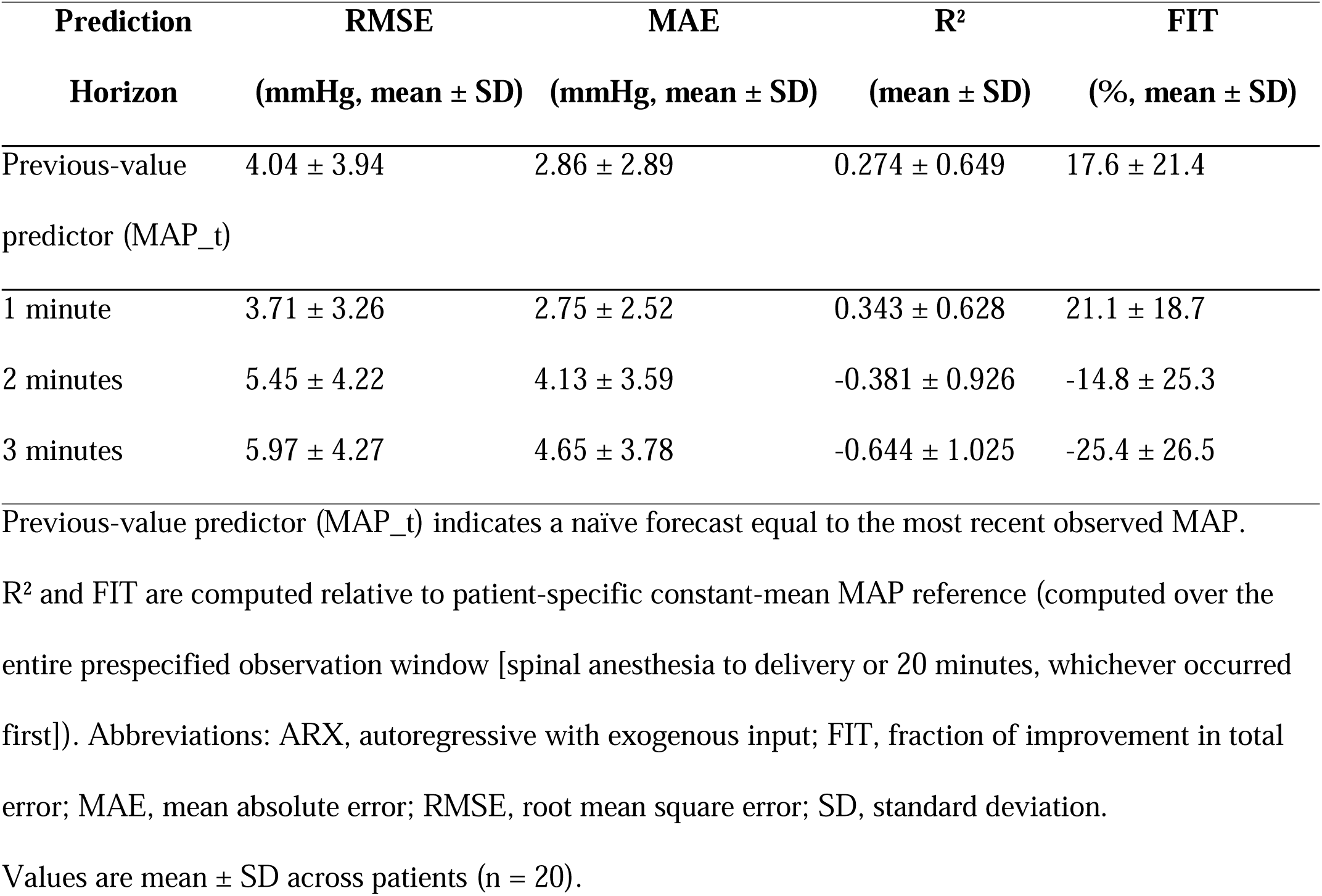
Patient-level predictive accuracy of ARX MAP forecasts at 1–3 minutes.

At the clinically relevant 1-minute prediction horizon, the mean (±SD) RMSE was 3.71 ± 3.26 mmHg, and the mean MAE was 2.75 ± 2.52 mmHg. The mean coefficient of determination (R²) was 0.343 ± 0.628, and the mean FIT was 21.1% ± 18.7% (Table 2).

Predictive performance decreased with increasing prediction horizon. At the 2-minute horizon, mean RMSE increased to 5.45 ± 4.22 mmHg, mean MAE increased to 4.13 ± 3.59 mmHg, and both R² (–0.381 ± 0.926) and FIT (–14.8% ± 25.3%) decreased, indicating worse predictive accuracy relative to the patient-specific total sum of squared deviations of the reference signal from its mean. Performance metrics at the 3-minute horizon exhibited a similar pattern (Table 2).

### Hypotension Prediction

The model’s ability to predict clinically significant hypotension (MAP <80% of baseline) is summarized in Supplemental Digital Content 3 and 4. At the 1-minute prediction horizon, GEE-estimated sensitivity was 57.39%, specificity 99.74%, PPV 86.16%, and NPV 97.84%. At this horizon, 17/398 (4.3%) evaluated time points met the hypotension definition (Supplemental Digital Content 3 and 4).

At 2 and 3 minutes, hypotension prediction performance declined substantially. GEE-estimated sensitivity decreased to 30.48% and 5.65%, respectively, although GEE-estimated specificity remained high (99.08% and 99.35%). PPV and NPV also decreased at longer horizons (2 minutes: PPV 62.50%, NPV 96.55%; 3 minutes: NPV 95.41%). At 3 minutes, PPV was not computed because all positive predictions originated from a single patient (Supplemental Digital Content 4).

### Clinical Outcomes Comparison

Exploratory comparisons of clinical outcomes between ARX-guided patients and matched nonconcurrent controls are presented in Table 3. Because these analyses used nonconcurrent controls and the study was not powered for clinical endpoints, all findings should be interpreted as hypothesis-generating.

**Table 3.**
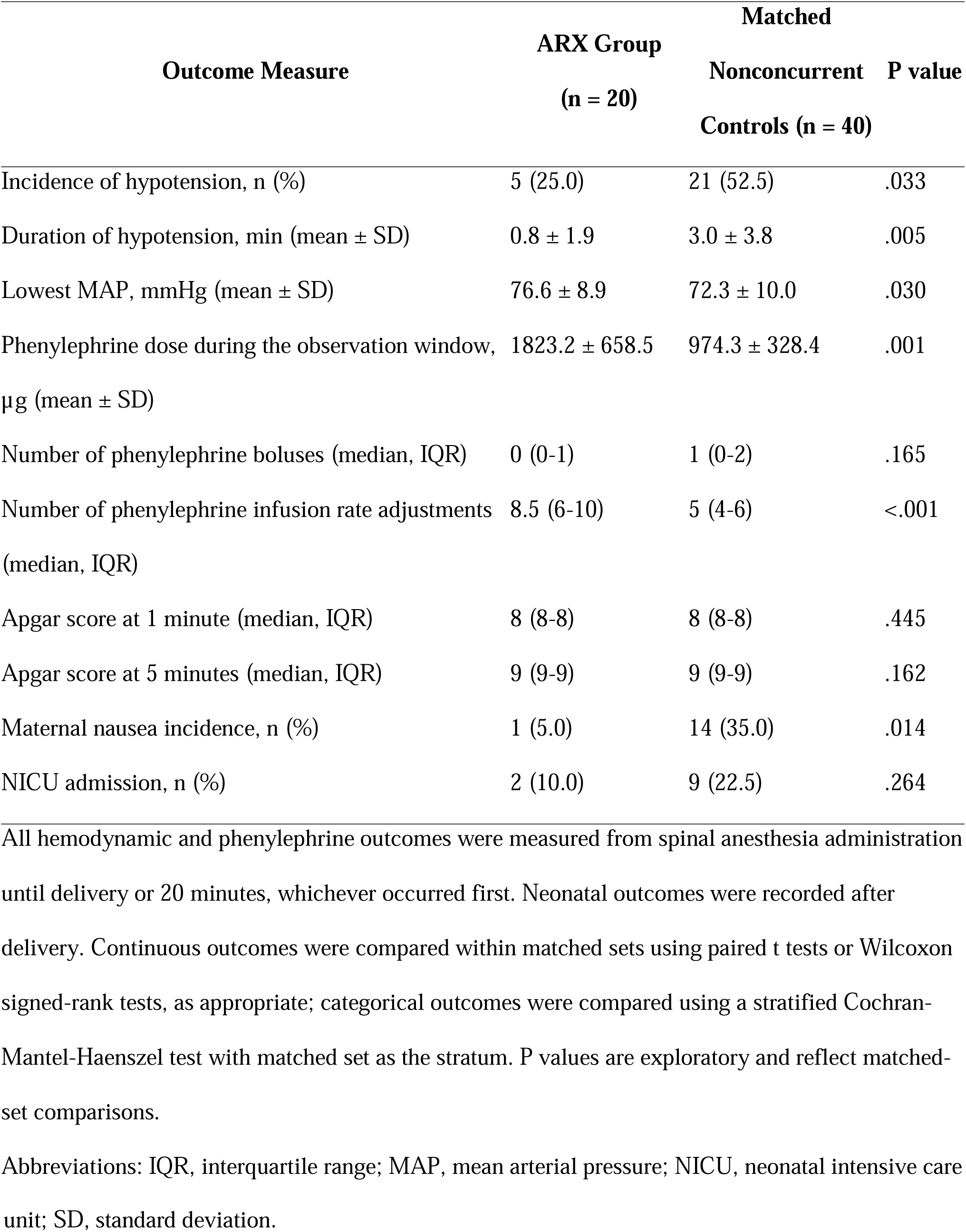
Exploratory outcomes in ARX-guided patients and matched nonconcurrent controls.

The average MAP and changes from baseline in both groups are shown in Figure 2. The incidence of hypotension during the prespecified observation window was lower in the ARX-guided group than in matched nonconcurrent controls (5/20 [25.0%] vs 21/40 [52.5%]; risk difference –27.5 percentage points; P = .033). The duration of hypotension was shorter in the ARX group (0.8 ± 1.9 minutes) compared with matched nonconcurrent controls (3.0 ± 3.8 minutes) (Supplemental Digital Content 5). Total phenylephrine dose during the observation window was higher in ARX-guided patients than in matched nonconcurrent controls (1823 ± 659 vs 974 ± 328 µg) (Supplemental Digital Content 6). The number of phenylephrine infusion rate adjustments in the ARX-guided patients was higher than in matched nonconcurrent controls (median 8.5 [IQR 6–10] vs 5 [IQR 4–6]; P < .001). Maternal nausea incidence was lower in the ARX-guided group (1/20 [5%]) compared with matched nonconcurrent controls (14/40 [35%]; P = .014). Neonatal outcomes were similar between groups, including median Apgar scores at 1 minute, which were 8 (IQR 8–8) in both groups (exploratory P = .445), and Apgar scores at 5 minutes, which were 9 (IQR 9–9) in both groups (P = .162).

**Figure 2.**
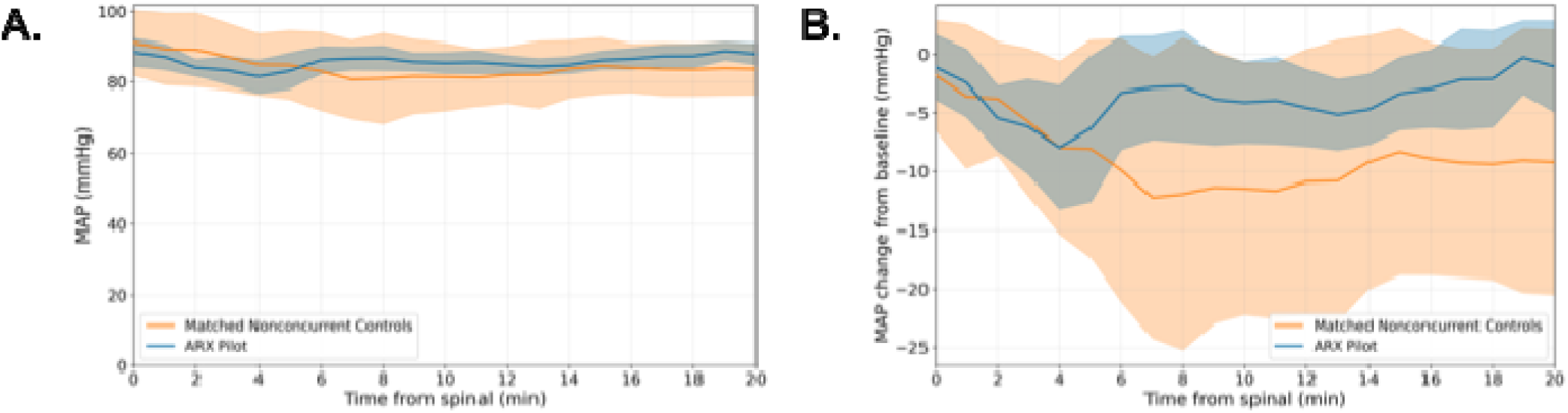
Mean (95% CI) MAP (A) and change from baseline MAP (B) by minute after spinal anesthesia in ARX-guided patients and matched nonconcurrent controls during the prespecified observation window (delivery or 20 minutes, whichever occurred first). The number of contributing patients may decrease over time due to delivery before 20 minutes. Baseline MAP was the mean of the last three pre-spinal NIBP measurements.

### Implementation

The ARX system displayed uninterrupted real-time forecasts in all 20 cases. One clinician-reported override occurred in 1 of 20 cases (5%), in which phenylephrine was administered despite a forecast of stable MAP.

## DISCUSSION

In this prospective, single-center pilot study of decision support during cesarean delivery under spinal anesthesia, an ARX model provided accurate 1-minute-ahead MAP forecasts (mean RMSE 3.71 mmHg) and demonstrated high specificity for impending hypotension at the 1-minute horizon; however, predictive performance deteriorated at 2–3 minutes (negative R² and FIT). In exploratory comparisons to matched nonconcurrent controls, ARX-guided care was associated with a shorter duration of hypotension during the prespecified observation window and less maternal nausea, while total phenylephrine dose and the number of infusion adjustments were higher. Neonatal outcomes (Apgar scores and NICU admission) were similar between groups, although this study was not powered to detect differences in neonatal endpoints, and the matched nonconcurrent control design limits causal inference.

One-minute accuracy (RMSE 3.71 ± 3.26 mmHg) was consistent with expectations for short-horizon forecasting from minute-by-minute noninvasive blood pressure measurements and with the retrospective model-development cohort (RMSE 3.6 ± 1.3 mmHg for 1-minute MAP prediction).[1] Although retrospective validation demonstrated acceptable performance up to 3 minutes,[1] 2–3-minute forecasts degraded in this prospective deployment. Several factors likely contributed to this degradation. First, forecasting beyond intermittently measured 1-minute MAP compounds measurement noise and reduces available information content. Second, time-varying hemodynamic responses to spinal sympathectomy and phenylephrine administration may be influenced by unmeasured factors, such as uterine displacement, fluid administration, patient anxiety, and surgical stimulation, that are not well captured by a low-order linear time-series model at longer horizons. Third, clinician responses to displayed forecasts may introduce a closed-loop interaction between predictions and phenylephrine dosing, reducing longer-horizon stability and potentially inducing dataset shift relative to the retrospective training cohort. These factors likely explain the higher 2- and 3-minute RMSE and FIT observed in the prospective setting compared with our original study.[1]

For hypotension prediction, the ARX system was highly specific with a high 1-minute NPV, but sensitivity was only moderate. This profile supports use as a clinical adjunct for early recognition of deteriorating trends with few false alarms. Because hypotension-positive minutes were uncommon in this pilot, PPV and NPV should be interpreted alongside event prevalence. Sensitivity may improve through hypotension-threshold recalibration, additional physiologic inputs (e.g., heart rate, trend features), or hybrid approaches combining time-series structure with nonlinear components.

Consensus guidance for cesarean delivery under spinal anesthesia emphasizes prophylactic vasopressor therapy and maintenance of arterial pressure close to baseline.[4] Definitions of hypotension vary widely, with relative thresholds such as <80% of baseline commonly used.[15] We defined hypotension as MAP <80% of baseline (institutional practice) and evaluated an ARX model trained to forecast MAP; although MAP and systolic pressure are related, different targets and definitions may affect apparent hypotension incidence and vasopressor dosing. Future work could assess whether forecasting systolic pressure (or converting MAP predictions to systolic targets) better aligns with consensus recommendations and clinician practice patterns.

Closed-loop vasopressor control is complementary. Computer-controlled phenylephrine infusions improved blood pressure control and reduced interventions versus manual titration,[8,9] and automated dual-vasopressor approaches reduced hypotension burden.[7] Our approach provides decision support rather than automated control, preserving clinician autonomy and potentially lowering implementation barriers (no direct pump control), but its effectiveness depends on clinician interpretation and action. Higher phenylephrine doses and more infusion adjustments in ARX-guided cases suggest forecasts may have encouraged more proactive therapy.

Predictive decision-support systems have also been evaluated outside obstetrics. Waveform-based machine-learning models can forecast hypotension 5–15 minutes ahead (e.g., Hypotension Prediction Index)[11] and, in adults undergoing noncardiac surgery, an early-warning system plus a treatment protocol reduced hypotension burden.[10] However, concerns remain about selection bias, generalizability, and overtreatment when algorithms prompt preemptive vasopressor therapy.[16] In obstetrics, invasive arterial waveforms are uncommon, although HPI performance has been evaluated with noninvasive finger-cuff waveforms.[12] While waveform-based approaches (e.g., HPI) have reported high sensitivity using invasive arterial pressure waveforms,[11] the ARX model achieves GEE-estimated 57.39% sensitivity using only intermittent NIBP readings, substantially lowering barriers to clinical implementation. The ARX approach is interpretable, uses routine intermittent NIBP with time-stamped phenylephrine inputs, and does not require pump integration, but its current utility appears limited to very short horizons (1 minute) and may still increase vasopressor exposure.

The exploratory clinical findings are biologically plausible: hypotension and blood pressure variability are associated with maternal nausea and vomiting during cesarean delivery, and prophylactic phenylephrine strategies that better maintain pressure improve maternal comfort.[17] Although we observed nausea in only one patient, which limits definitive conclusions, this finding aligns with the expected reduction in hypotension burden.

Higher phenylephrine exposure may carry trade-offs, including bradycardia and potential reductions in maternal cardiac output, motivating interest in norepinephrine; however, evidence remains insufficient to establish clear superiority for norepinephrine in this setting.[18] Future ARX trials should prespecify and report bradycardia, reactive hypertension, and cardiac output surrogates, alongside hypotension metrics, to ensure a balanced safety/benefit assessment.

Baseline selection is also important. Operating-room “baseline” blood pressure may exceed preoperative ambulatory measurements, potentially increasing vasopressor use if targets are tied rigidly to that baseline.[5,13] Because ARX forecasting is coupled to clinician titration, standardizing and explicitly reporting baseline definition will be critical for comparisons across sites and for interpreting phenylephrine dosing differences.

### Limitations

Limitations include small sample size and limited power for clinical outcomes (comparisons are hypothesis-generating), open-label design with potential Hawthorne effects, and nonconcurrent controls; despite matching on attending anesthesiologist and intrathecal bupivacaine dose, unmeasured confounding and temporal practice changes may remain. Imputation of rare missing NIBP measurements in matched nonconcurrent controls could affect hypotension-duration estimates, although missingness was low. Although matched-set methods were used for exploratory comparisons, the pilot sample size limited precision, and analyses were not adjusted for multiple comparisons; future randomized studies should emphasize effect sizes with confidence intervals and prespecified safety endpoints.

### Implications and future directions

These results support the technical operability of real-time MAP forecasting in the obstetric operating room and provide preliminary effect-size estimates for a definitive trial. Next steps include prospective randomized evaluation against concurrent controls, prespecified targets aligned to consensus guidance, broader safety reporting (hypertension, bradycardia, interventions for nausea/vomiting), model enhancement with additional physiologic and therapeutic inputs, and multicenter validation to test generalizability across monitors, practice patterns, and patient populations.

In summary, an ARX decision-support system provided accurate 1-minute-ahead MAP forecasts during cesarean delivery under spinal anesthesia and was associated with reduced hypotension duration with increased phenylephrine use in exploratory comparisons to matched nonconcurrent controls. Larger randomized studies are warranted to confirm efficacy, characterize trade-offs, and define optimal integration into clinical practice.

## Supporting information

Supplemental Digital Content

## Data Availability

In accordance with the Mass General Brigham Institutional Review Board requirements, the research data supporting this project may not be publicly shared.

## Clinical trial number

Not applicable (single-arm prospective pilot; no assignment to treatment groups).

## Prior Presentations

None

## Generative AI use

During the preparation of this manuscript, the authors used a large language model (ChatGPT, OpenAI) for language editing and to improve clarity. The authors reviewed and edited all AI-assisted text and take full responsibility for the content.

## Acknowledgments

The authors are deeply grateful to Erin McKenna, MBA, MS; Kalpana Kamath, PhD; Chun Leung Ng, PhD; and Glenn Miller, PhD (Mass General Brigham, Boston, MA) for their sustained strategic guidance and programmatic and operational support over many years that helped enable this work.

## Abbreviations

ARX: autoregressive with exogenous input
BMI: body mass index
FIT: fraction of improvement in total error
GEE: generalized estimating equations
IRB: Institutional Review Board
IQR: interquartile range
MAE: mean absolute error
MAP: mean arterial pressure
NIBP: noninvasive blood pressure
NICU: neonatal intensive care unit
NPV: negative predictive value
PPV: positive predictive value
RMSE: root mean square error
SD: standard deviation
SMD: standardized mean difference

## REFERENCES

[1] S.C. Davoud, B. Ozaslan, E.M. Aiello, R. Kleinlein, B. Eberhard, H. Hassan, F.J. Doyle, V.P. Kovacheva, Empirical pharmacodynamic model of phenylephrine and intrathecal bupivacaine for mean arterial pressure prediction in obstetric patients presenting for elective cesarean delivery under spinal anesthesia, J Clin Monit Comput (2025). 10.1007/s10877-025-01288-w.

[2] A.S. Habib, A review of the impact of phenylephrine administration on maternal hemodynamics and maternal and neonatal outcomes in women undergoing cesarean delivery under spinal anesthesia, Anesth Analg 114 (2012) 377–390. 10.1213/ANE.0b013e3182373a3e.

[3] W.D. Ngan Kee, K.S. Khaw, P.E. Tan, F.F. Ng, M.K. Karmakar, Placental transfer and fetal metabolic effects of phenylephrine and ephedrine during spinal anesthesia for cesarean delivery, Anesthesiology 111 (2009) 506–512. 10.1097/ALN.0b013e3181b160a3.

[4] S.M. Kinsella, B. Carvalho, R.A. Dyer, R. Fernando, N. McDonnell, F.J. Mercier, A. Palanisamy, A.T.H. Sia, M. Van de Velde, A. Vercueil. International consensus statement on the management of hypotension with vasopressors during caesarean section under spinal anaesthesia, Anaesthesia 73 (2018) 71–92. 10.1111/anae.14080.

[5] W.D. Ngan Kee, K.S. Khaw, F.F. Ng, B.B. Lee, Prophylactic phenylephrine infusion for preventing hypotension during spinal anesthesia for cesarean delivery, Anesthesia and Analgesia 98 (2004) 815–21. 10.1213/01.ane.0000099782.78002.30.

[6] R.A. Dyer, A.R. Reed, D. van Dyk, M.J. Arcache, O. Hodges, C.J. Lombard, J. Greenwood, M.F. James, Hemodynamic effects of ephedrine, phenylephrine, and the coadministration of phenylephrine with oxytocin during spinal anesthesia for elective cesarean delivery, Anesthesiology 111 (2009) 753–65. 10.1097/ALN.0b013e3181b437e0.

[7] B.L. Sng, H.S. Tan, A.T.H. Sia, Closed-loop double-vasopressor automated system vs manual bolus vasopressor to treat hypotension during spinal anaesthesia for caesarean section: a randomised controlled trial, Anaesthesia 69 (2014) 37–45. 10.1111/anae.12460.

[8] W.D. Ngan Kee, Y.H. Tam, K.S. Khaw, F.F. Ng, L.A. Critchley, M.K. Karmakar, Closed-loop feedback computer-controlled infusion of phenylephrine for maintaining blood pressure during spinal anaesthesia for caesarean section: a preliminary descriptive study, Anaesthesia 62 (2007) 1251–1256. 10.1111/j.1365-2044.2007.05257.x.

[9] W.D. Ngan Kee, K.S. Khaw, F.F. Ng, Y.H. Tam, Randomized comparison of closed-loop feedback computer-controlled with manual-controlled infusion of phenylephrine for maintaining arterial pressure during spinal anaesthesia for caesarean delivery, Br J Anaesth 110 (2013) 59–65. 10.1093/bja/aes339.

[10] M. Wijnberge, B.F. Geerts, L. Hol, N. Lemmers, M.P. Mulder, P. Berge, J. Schenk, L.E. Terwindt, M.W. Hollmann, A.P. Vlaar, D.P. Veelo, Effect of a Machine Learning-Derived Early Warning System for Intraoperative Hypotension vs Standard Care on Depth and Duration of Intraoperative Hypotension During Elective Noncardiac Surgery: The HYPE Randomized Clinical Trial, JAMA 323 (2020) 1052–1060. 10.1001/jama.2020.0592.

[11] F. Hatib, Z. Jian, S. Buddi, C. Lee, J. Settels, K. Sibert, J. Rinehart, M. Cannesson, Machine-learning Algorithm to Predict Hypotension Based on High-fidelity Arterial Pressure Waveform Analysis, Anesthesiology (2018). 10.1097/ALN.0000000000002300.

[12] L. Frassanito, C. Sonnino, A. Piersanti, B.A. Zanfini, S. Catarci, P.P. Giuri, M. Scorzoni, G.L. Gonnella, M. Antonelli, G. Draisci, Performance of the Hypotension Prediction Index With Noninvasive Arterial Pressure Waveforms in Awake Cesarean Delivery Patients Under Spinal Anesthesia, Anesth Analg 134 (2022) 633–643. 10.1213/ANE.0000000000005754.

[13] D.G.P. Luther, S. Scholes, N. Wharton, S.M. Kinsella, Selection of baseline blood pressure to guide management of hypotension during spinal anaesthesia for caesarean section, International Journal of Obstetric Anesthesia 45 (2021) 130–132. 10.1016/j.ijoa.2020.11.010.

[14] V.P. Kovacheva, W. Armero, G. Zhou, D. Bishop, R. Dyer, B. Carvalho, Investigation of the Optimum Baseline Blood Pressure for Spinal Anesthesia to Guide Vasopressor Management for Elective Cesarean Delivery: A Case-Control Design, Cureus 15 (2023) e45380. 10.7759/cureus.45380.

[15] S. Klohr, R. Roth, T. Hofmann, R. Rossaint, M. Heesen, Definitions of hypotension after spinal anaesthesia for caesarean section: literature search and application to parturients, Acta Anaesthesiol Scand 54 (2010) 909–21. 10.1111/j.1399-6576.2010.02239.x.

[16] F. Michard, E. Futier, Predicting intraoperative hypotension: from hope to hype and back to reality, Br J Anaesth 131 (2023) 199–201. 10.1016/j.bja.2023.02.029.

[17] J.E. Lee, R.B. George, A.S. Habib, Spinal-induced hypotension: Incidence, mechanisms, prophylaxis, and management: Summarizing 20 years of research, Best Practice & Research Clinical Anaesthesiology 31 (2017) 57–68. 10.1016/j.bpa.2017.01.001.

[18] M. Heesen, N. Hilber, K. Rijs, R. Rossaint, T. Girard, F.J. Mercier, M. Klimek, A systematic review of phenylephrine vs. noradrenaline for the management of hypotension associated with neuraxial anaesthesia in women undergoing caesarean section, Anaesthesia 75 (2020) 800–808. 10.1111/anae.14976.

