## Supplemental Digital Content for "Autoregressive With Exogenous Input (ARX) Decision Support for Blood Pressure Maintenance During Cesarean Delivery Under Spinal Anesthesia: A Prospective Pilot Study With Matched Nonconcurrent Controls"

**SUPPLEMENTAL DIGITAL CONTENT 1**

SUPPLEMENTAL METHODS:

To allow for a real-time interaction (input and display) with the previously developed ARX model used to predict MAP for up to a 3-minute horizon, we developed a simple Python user interface using the built-in Tkinter library. The ARX model used for real-time prediction has been previously described and validated.[1] Tkinter was selected for its minimal dependencies, cross-platform compatibility, and simplicity. Additionally, this interface could be executed on any system capable of running a standard Python interpreter, without requiring internet connectivity or specialized hardware, thereby supporting its deployment in resource-constrained clinical settings. The interface accepted time-stamped inputs (MAP and phenylephrine infusion/bolus dosing) and the intrathecal bupivacaine dose and time of administration, and generated 1- to 3-minute-ahead MAP forecasts. An example of the user interface screen is shown in Supplemental Figure S1. The workflow included in-person research staff (M.N., S.D., or V.K.) who manually entered MAP values from the anesthesia monitor at 1-minute intervals and recorded phenylephrine dosing changes and the intrathecal bupivacaine dose/time as they occurred. Medications were administered by the anesthesia team according to institutional practice; research staff did not administer study medications.

**Supplemental Figure S1.** Representative screenshot of the ARX decision-support user interface (Python/Tkinter) used for real-time entry of time-stamped MAP and phenylephrine/bupivacaine inputs and display of 1–3-minute-ahead MAP forecasts; the display provided no dosing recommendations.

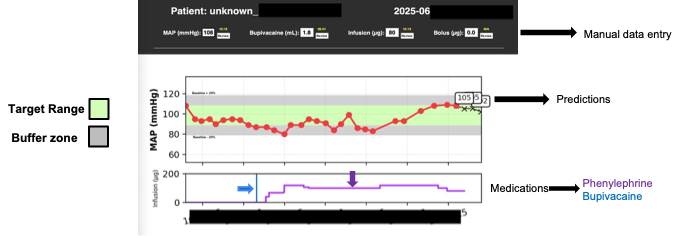

**SUPPLEMENTAL DIGITAL CONTENT 2**

Institutional vasopressor administration protocol.

Protocol for Management of Maternal Blood Pressure in Normotensive, Non-Preeclamptic Patients During Cesarean Delivery

1. Place ECG leads, NIBP cuff, and pulse oximeter ≥5 min before spinal, and set NIBP to cycle every 1 min.

2. Administer ondansetron 4 mg IV prior to spinal anesthesia.

3. Start prophylactic phenylephrine infusion at 20-40 µg/min at the time of spinal.

4. If MAP decreases by ≥20% from baseline or heart rate increases to over 100 beats/min, administer bolus 80-120 µg phenylephrine.

5. If MAP is still low, administer another bolus of 80-120 µg of phenylephrine, and consider increasing the infusion rate by 40 µg/min.

6. Avoid using ephedrine unless the heart rate is <50 beats/min and MAP is low after giving at least 2 boluses of phenylephrine.

7. Reserve glycopyrrolate or atropine for heart rate ≤50 beats/min.

**SUPPLEMENTAL DIGITAL CONTENT 3**

**Aggregated confusion matrices**

For each prediction horizon considered by the ARX model (1, 2, and 3 minutes ahead), we report the aggregated confusion matrices (Supplemental Table S1) associated with the binary classification task of anticipating an upcoming hypotensive event. At each minute and for each patient in the study group, the continuous blood pressure forecast was converted into a binary label indicating whether a hypotensive episode was predicted to occur within the corresponding forecast horizon, using the same clinically defined threshold applied throughout the study. Confusion matrices were first computed at the individual-patient level and subsequently aggregated across all patients.

| **Supplemental Table S1. Aggregated confusion matrices for ARX prediction of hypotension (MAP <80% of baseline) at 1-, 2-, and 3-minute horizons during the prespecified observation window in the ARX-guided cohort.** | | |
| --- | --- | --- |
| **Prediction horizon: 1 min (n = 398 time points)** |  |  |
| **Ground truth/Prediction** | **No hypotension** | **Hypotension** |
| No hypotension | 380 | 1 |
| Hypotension | 8 | 9 |
| **Prediction horizon: 2 min (n=378 time points)** |  |  |
| **Ground truth/Prediction** | **No hypotension** | **Hypotension** |
| No hypotension | 358 | 3 |
| Hypotension | 12 | 5 |
| **Prediction horizon: 3 min (n=358 time points)** |  |  |
| **Ground truth/Prediction** | **No hypotension** | **Hypotension** |
| No hypotension | 339 | 2 |
| Hypotension | 16 | 1 |
| Counts represent minute-level time points pooled across patients (n=398, 378, and 358 evaluable time points at 1, 2, and 3 minutes, respectively). | | |

**SUPPLEMENTAL DIGITAL CONTENT 4**

| **Supplemental Table S2. GEE-estimated sensitivity, specificity, PPV, and NPV with 95% CIs for predicting hypotension events at specified horizons.** | | | | |
| --- | --- | --- | --- | --- |
| **Prediction Horizon** | **Sensitivity (%)** | **Specificity (%)** | **PPV (%)** | **NPV (%)** |
| Previous-value predictor (MAP_t) | 58.46 [54.47, 62.35] | 98.39 [96.24, 99.32] | 66.55 [65.62, 67.48] | 98.03 [94.93, 99.25] |
| 1 minute | 57.39 [53.40, 61.29] | 99.74 [98.24, 99.96] | 86.16 [39.13, 98.37] | 97.84 [94.74, 99.13] |
| 2 minutes | 30.48 [14.68, 52.75] | 99.08 [96.34, 99.78] | 62.50 [44.32, 77.73] | 96.55 [91.50, 98.64] |
| 3 minutes | 5.65 [0.87, 29.01] | 99.35 [95.69, 99.91] | * | 95.41 [88.33, 98.28] |
| Previous-value predictor (MAP_t) indicates a naïve forecast equal to the most recent observed MAP.  Note: Performance metrics in Supplemental Table S2 are GEE-estimated marginal (population-average) measures that account for within-patient clustering and may differ from simple ratios computed directly from pooled confusion-matrix cell counts in Supplemental Table S1.  Abbreviations: MAP, mean arterial pressure; NPV, negative predictive value; PPV, positive predictive value. *PPV and confidence intervals were not computed in this case because all positive predictions originated from a single patient, precluding variance estimation under a clustered GEE framework*.* | | | | |

**SUPPLEMENTAL DIGITAL CONTENT 5**

**Supplemental Figure S2.** Distribution of hypotension duration during the observation window in ARX-guided patients and matched nonconcurrent controls.

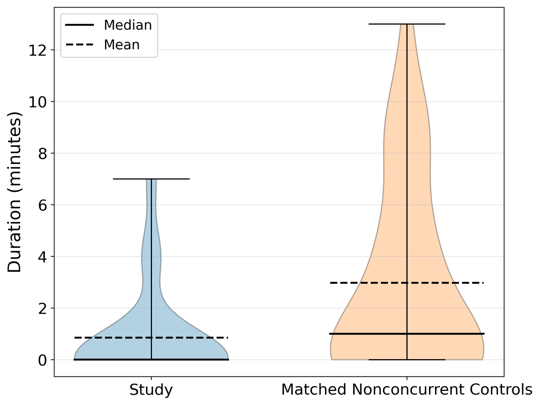

**SUPPLEMENTAL DIGITAL CONTENT 6**

**Supplemental Figure S3.** Distribution of phenylephrine dose during the observation window in ARX-guided patients and matched nonconcurrent controls.

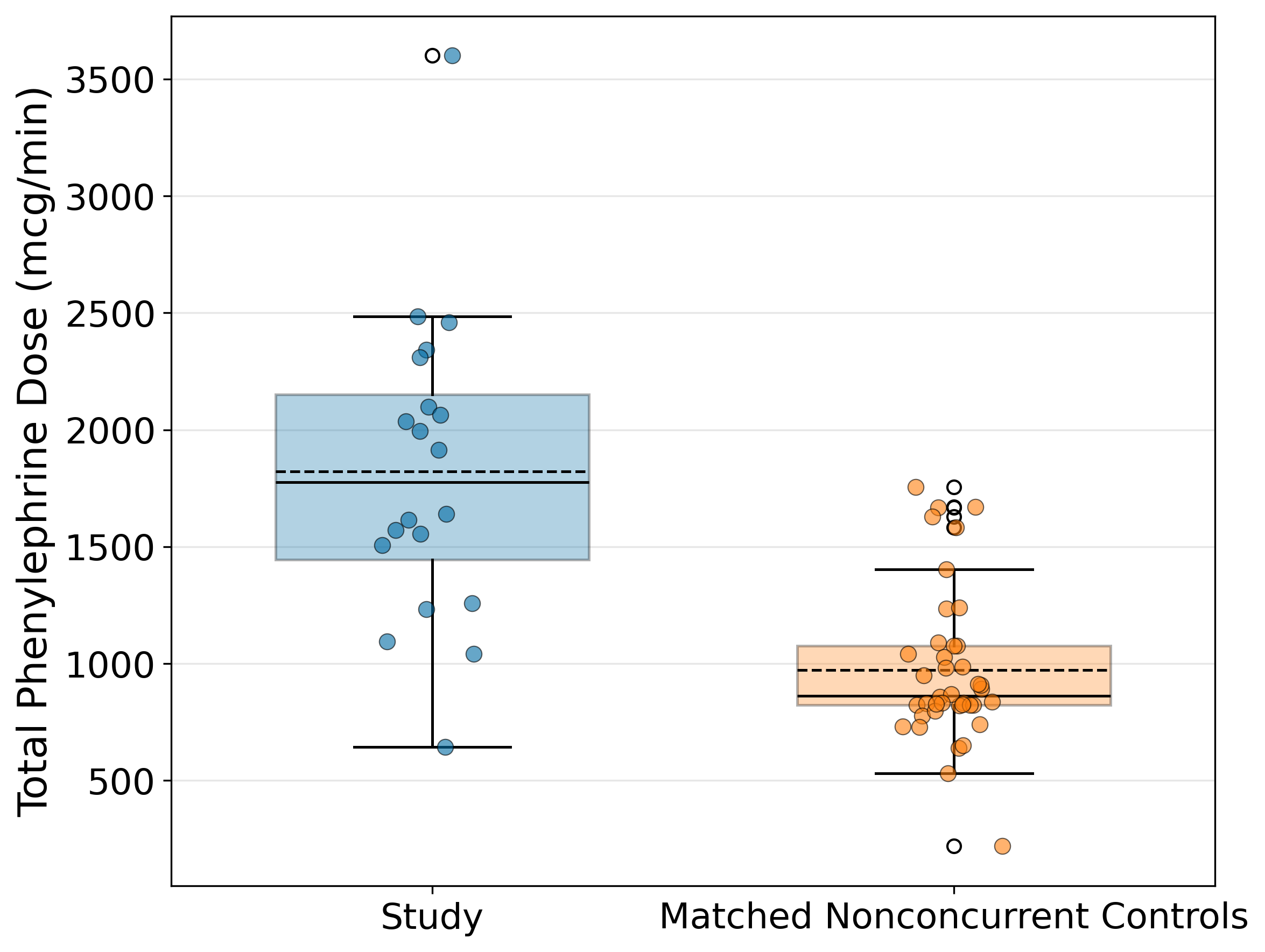
